# Physical rehabilitation versus no physical rehabilitation after total hip and knee arthroplasty: A replication trial in 169 patients with a 12-month follow-up (DRAW2)

**DOI:** 10.1101/2025.09.02.25334893

**Authors:** T. Mark-Christensen, K. Thorborg, T. Kallemose, T. Bandholm

## Abstract

**Importance:** Although physical rehabilitation is widely recommended after total hip (THA) and knee (TKA) arthroplasty, its fundamental clinical effectiveness—defined as the added benefit beyond natural recovery—remains uncertain. We recently published the DRAW1 trial and did not find physical rehabilitation to be superior to no physical rehabilitation following THA and TKA. This was a challenging finding, and its validity was questioned by the rehabilitation community, so we decided to replicate the DRAW1 trial using the same trial design and site.

**Objective:** To replicate the DRAW1 trial by comparing the effectiveness of 6-weeks of home-based <u>tele</u>rehabilitation, home-based rehabilitation or no physical rehabilitation following THA and TKA in terms of self-reported function.

**Design:** 3-arm parallel-group randomized, controlled, replication trial with blinded outcome assessments. 479 patients (221 THA/258 TKA) were screened for eligibility before the targeted sample size was reached. Following discharge, 52 patients were randomized to home-based telerehabilitation, 58 to home-based rehabilitation, and 59 to no physical rehabilitation for 6 weeks. Outcome measures were assessed blinded in an outpatient-setting at baseline (post-discharge), at the end of intervention (6 weeks – primary endpoint), and 3 and 12 months postoperatively. The primary outcome was the Hip disability and Osteoarthritis Outcome Score (HOOS)/ Knee injury and Osteoarthritis Outcome Score (KOOS)-subscale: function in daily living (ADL).

**Results:** In the primary intention-to-treat analysis—comparing physical rehabilitation (home-based telerehabilitation and home-based rehabilitation) to no physical rehabilitation—the mean group-differences for the primary outcome were −0.5 (95%CI: −3.1 to 2.1; p = 0.70) KOOS/HOOS points at 6 weeks (primary endpoint; MCID=10 KOOS/HOOS points), and 0.8 (95%CI: −1.7 to 3.4; p = 0.52) and 0.4 (95%CI: −2.2 to 2.9; p = 0.48) points at the 3- and 12-months follow-ups, respectively.

**Conclusion:** The main finding from the DRAW1 trial was replicated: once again we did not find physical rehabilitation to be superior to no physical rehabilitation following THA or TKA in terms of self-reported function. Thus, the fundamental clinical effectiveness of physical rehabilitation in this clinical context could not be established.

## INTRODUCTION

Exercise-based rehabilitation is commonly prescribed following hip- and knee arthroplasty (THA and TKA) for osteoarthritis, to enhance postoperative recovery ^1,2^. Clinical practice varies widely, ranging from supervised rehabilitation in outpatient centers to brief instruction followed by unsupervised home exercise ^3,4^. Despite its widespread use, no specific rehabilitation strategy has shown consistent superiority across systematic reviews ^5–8^, raising questions about whether physical rehabilitation following THA and TKA provides benefit beyond natural recovery, i.e., its fundamental clinical effectiveness. This aligns with the principles of the international Choosing Wisely campaign, which promotes reducing low-value or potentially unnecessary care ^9^. In rehabilitation, this means questioning routine provision of structured physical rehabilitation for all patients after THA or TKA, particularly when robust evidence of benefit is lacking.

To investigate this, we conducted systematic reviews comparing postoperative rehabilitation to no rehabilitation following THA and TKA ^10,11^. However, the limited number of trials (5 trials with 314 participants) and their heterogeneity prevented any conclusion to be drawn - and thus highlighted the need for a pragmatic, adequately powered randomized controlled trial with real-world applicability. As a consequence, we designed the pragmatic DRAW1 trial to evaluate the fundamental effectiveness of exercise-based rehabilitation in a mixed THA and TKA population ^12^. We did not find exercise-based rehabilitation to be superior to no rehabilitation in terms of self-reported function or secondary outcomes ^13^. These findings challenged our own as well as the rehabilitation community’s pre-conceptions because post-surgical rehabilitation is an established clinical standard in many healthcare systems and widely regarded as essential for optimizing recovery (beyond natural recovery). Given the potentially practice-changing nature of the DRAW1 trial findings ^13^, the DRAW2 trial was designed as a direct replication using the same protocol, setting, and design.

### Objective

To replicate the DRAW1 trial by comparing physical rehabilitation (combined intervention groups) to no rehabilitation following THA or TKA, using the same superiority hypothesis.

## METHOD

This is the primary trial report for the DRAW2 replication trial (“**D**oes **R**ehabilitation after total hip and knee **A**rthroplasty “**W**ork 2”?). The DRAW2 trial was designed as a replication trial of the DRAW1 trial using the DRAW1 trial protocol ^13^ and study site.

As with DRAW1, the DRAW2 trial used a superiority, three-arm, parallel-group, randomized, controlled, trial (RCT) design with blinded outcome assessments at baseline (before intervention), at 6 weeks (end of intervention, primary endpoint), and follow-ups at 3 and 12 months. The DRAW-trial protocol was based on the PREPARE trial guide ^14^ and the SPIRIT checklist ^15^ and published open access ^12^. The DRAW2 trial was conducted using the identical protocol as DRAW1, with one predefined deviation: baseline assessments were allowed to be split over two visits when deemed necessary by the assessing physiotherapist. The DRAW2 trial report adheres to the Consolidated Standards of Reporting Trials (CONSORT) ^16^ and was pre-registered on July 2. 2021 at ClinicalTrials.gov (NCT04960241) after approval from the Ethics Committee of the Capital Region Denmark (ID: H-18056678) ^17^. The procedures followed were in accordance with the ethical standards of the responsible committee on human experimentation (institutional and national) and with the Helsinki Declaration of 1975, as revised in 2000 ^18^. A full trial protocol for the DRAW1 trial and the current replication (DRAW2) trial was published open access on February 25. 2021 ^12^. Below we summarize the protocol. For details, please see the protocol ^12^.

### Participants

To reflect current clinical practice and to increase external validity and generalizability, few eligibility criteria were used.

The inclusion criteria were:

- Primary, unilateral THA or TKA due to osteoarthritis.
- Referred to receive postoperative rehabilitation at our institution (usual practice in the Capital Region of Denmark).
- Able to speak, read, and understand Danish language.
- Aged 18 years or more.

The exclusion criteria were:

- Not able to comply with exercise instructions.
- Discharged to a nursing home facility or receiving in-home rehabilitation by home care.

### Setting

All patients were recruited consecutively from three outpatient rehabilitation centers on the isle of Bornholm, Denmark. All had undergone unilateral primary THA or TKA and were referred to municipal, free-of-charge postoperative rehabilitation—reflecting usual care in Eastern Denmark. Prior to discharge, patients had achieved independent basic mobility and manageable pain levels. They were invited by letter to initiate rehabilitation approximately five to seven days after discharge. At the first consultation, an experienced physiotherapist provided oral and written trial information, obtained informed consent, and conducted baseline assessments prior to randomization. If consent was not given, patients were offered usual care (home-based rehabilitation). Randomization was performed by an external physiotherapist using a computer-generated sequence (1:1:1), concealed in opaque, sealed envelopes. Block randomization (block size 9) ensured balance due to a limited number of telerehabilitation units.

#### Interventions

The trial compared three post-discharge strategies: home-based telerehabilitation, home-based rehabilitation, and no physical rehabilitation, in a mixed population of THA and TKA patients. The study was originally initiated following a request from the municipal rehabilitation service to evaluate a telerehabilitation solution against usual care (home-based rehabilitation). As exercise modality and level of supervision are not known confounders, the two rehabilitation arms were combined in the primary analysis. This approach, first applied in DRAW1, was retained in DRAW2. All interventions were initiated in the first or second visit during outpatient consultations following discharge, all within two weeks of the primary surgery.

### Home-based telerehabilitation

Patients randomized to this intervention received interactive virtual rehabilitation via a mobile app using ICURA sensor technology (www.icura.dk). The system combines motion sensors that monitor exercise quality and quantity with real-time visual feedback through the app. A key feature is that physiotherapists can remotely supervise and adjust patient adherence and progression. The ICURA system is already in use across several Danish rehabilitation settings and reflects current clinical practice (picture 1).

### Home-based rehabilitation

Patients in this group followed the same exercise program as the telerehabilitation group but received only a written version, without technological support. This reflects usual care. The program was developed using Exorlive templates (www.exorlive.com) and included links to short instructional videos for each exercise (picture 2).

### No physical rehabilitation

Patients in this group received no prescribed therapeutic exercise or follow-up beyond advice to resume activities of daily living (e.g., walking, vacuuming) at their own pace. The rationale for this approach and terminology is detailed in the published trial protocol ^12^.

All participants received identical pamphlets with information about their surgery, expected discomforts, complications, and advice on returning to daily activities. For the two rehabilitation groups, exercise instructions were included as add-ons. Each program comprised four standardized exercises tailored to THA or TKA, performed daily (3 sets of 10 repetitions) at 15 RM intensity, progressing biweekly. This protocol reflects usual care at our institution. Full intervention details following TIDieR ^19^ and CERT guidelines ^20^ are publicly available via Harvard Dataverse ^21^.

#### Outcome

Licensed physiotherapists trained by the primary investigator conducted all outcome assessments using standardized protocols. Data were collected at baseline (first consultation), 6 weeks (end of intervention), 3 months, and 12 months postoperatively. **The primary outcome** was the between-group difference in the mean HOOS/KOOS ADL subscale score at 6 weeks ^22,23^. This subscale was selected based on patient input and its clinical relevance to daily functioning, the target of the rehabilitation effort. Due to their similar psychometric properties, HOOS and KOOS ADL scores were pooled for analysis. **Secondary outcomes** included between-group differences at all follow-ups in HOOS/KOOS subscales for pain, symptoms, and quality of life. Secondary outcomes further included a patient global assessment of function (0–100 VAS), the 30-s chair stand test, and the 4×10 m walk test, as recommended by OARSI ^24,25^. Patient satisfaction and exercise adherence (0–100 %) were recorded at 6 weeks. Additional measures included use of analgesics and walking aids, adverse events, and physiotherapist time use across groups.

All outcomes were documented in medical records and transferred in collaboration with the assessing physiotherapist.

#### Blinding

Given the nature of the interventions, participants and care providers could not be blinded to group allocation, but they were blinded to the trial hypothesis to reduce expectation and response bias ^26,27^. Outcome assessors were blinded to allocation at all time points, with baseline assessments conducted before randomization. Patients were instructed not to reveal their allocation, and assessors were trained to avoid cues or questions that could unblind them. The principal investigator, who maintained contact with participants, was not involved in outcome assessments and was aware of allocations.

#### Sample size

Sample size estimates mirrored those from the DRAW1 trial to ensure methodological comparability, using a 10-point superiority margin on the HOOS/KOOS ADL subscale (SD=20, α=0.05, and power=0.80). To account for an expected 10 % loss to follow-up ^28^, we calculated with 56 patients in each group, resulting in a total of 168 patients (3 x 56 = 168).

#### Statistical methods

A detailed statistical analysis plan was published as part of the trial protocol for the DRAW1 trial ^12^ and made publicly available prior to DRAW2 data analysis. The primary analysis aimed to test if the mean difference on the HOOS/KOOS ADL subscale score at 6 weeks for physical rehabilitation (home-based telerehabilitation and home-based rehabilitation) was superior to no physical rehabilitation (primary trial objective). This was done by independent two-sample t-tests or Wilcoxon sum rank-test, if the data could not be assumed to be normally distributed. Comparison of the two physical rehabilitation interventions individually to the no physical rehabilitation intervention were analyzed the same way as the primary analysis. Analysis was also repeated for 3- and 12-month follow-up times. Results from the analysis are presented as differences in mean changes (baseline to follow-ups) with two-sided 95% confidence intervals (95%CI) and p-values for superiority of physical rehabilitation. Chi-squared test was used for comparisons of nominal outcomes. All analyses were performed as intention-to-treat (analyzed as randomized). Normality assumptions were evaluated by quantile-quantile plots and histograms. Missing data were imputed using multiple imputations were used to impute missing data, with models fitted for each variable including type of surgery, age, gender and prior assessments for the specific variable being imputed. Additionally, a per protocol analysis was conducted for the primary outcome. All patients having an exercise adherence of at least 80 % were considered adherent and compared to patients in the ‘no physical rehabilitation’ group. Adjustment for multiple testing using Bonferroni correction was done for secondary analyses. P-values of < 0.05 were considered statically significant. Imputation of missing data was performed in R 4.3.2 ^29^ and remaining analysis in STATA version 16.1 ^30^.

## RESULTS

Participant flow: 479 patients (221 THA / 258 TKA) were referred to our outpatient rehabilitation institution between the 29^th^ of March 2021 to the 22^nd^ of March 2024 and assessed for eligibility. 93 patients were not eligible, and 217 patients declined participation, leaving 169 participants to be included (inclusion rate: 43.8 %) (Figure 1). Baseline assessments were performed over two visits for 76 participants (45 %).

**Picture 1.**
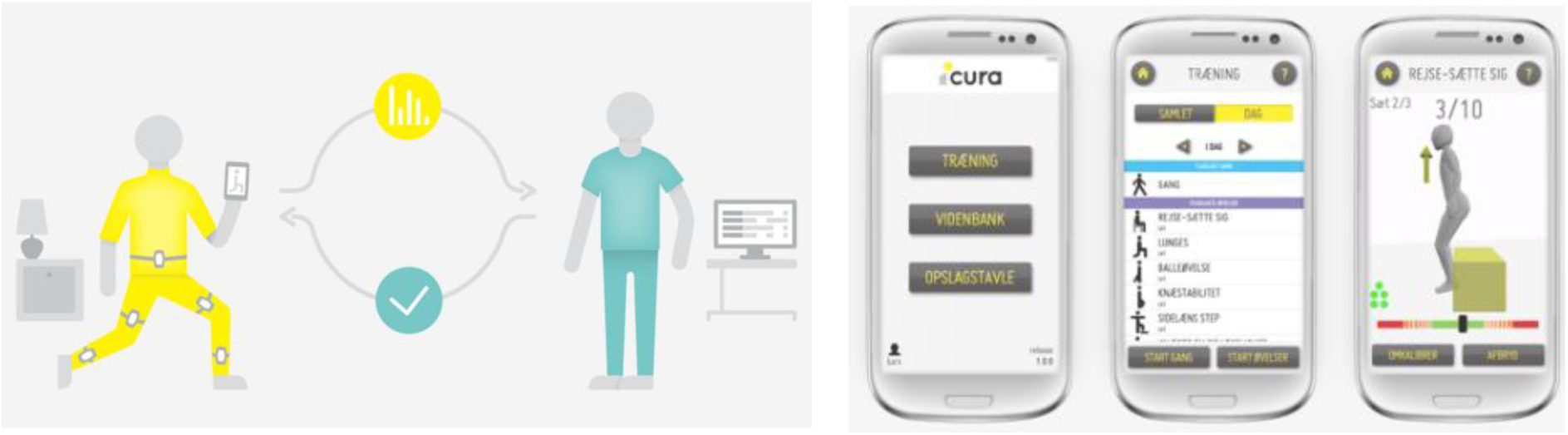
Features of home-based telerehabilitation (ICURA).

**Picture 2.**
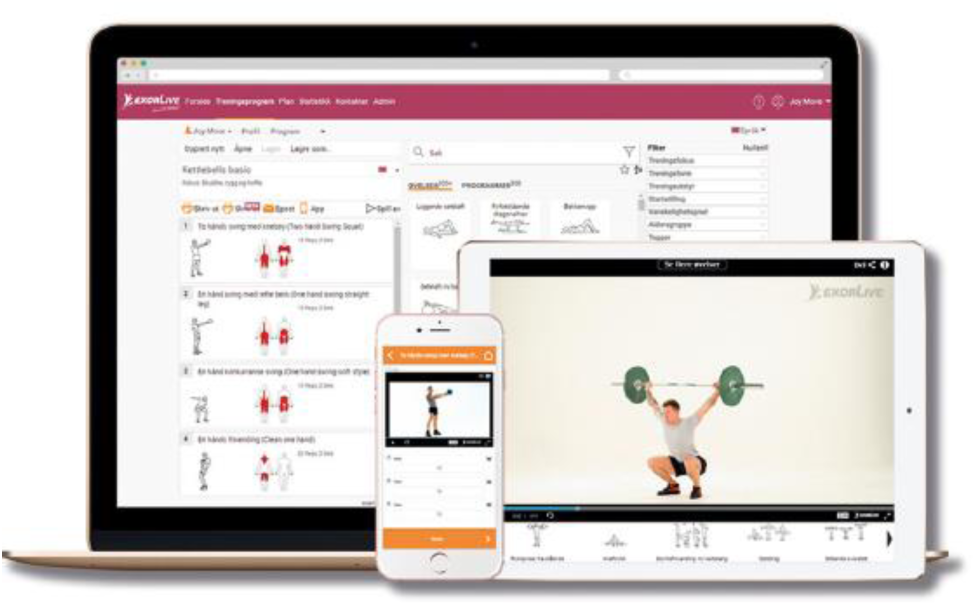
Home-based rehabilitation (Exorlive)

### Lost to follow-up

15 patients were lost before the first follow-up at the end of the 6-week intervention period (five from the home-based telerehabilitation intervention, four from the home-based rehabilitation intervention, and six from the no physical rehabilitation intervention). Total loss to follow-up during the entire trial period was 34 patients (20 %) (nine from the home-based telerehabilitation intervention, nine from the home-based rehabilitation intervention, and 16 from the no physical rehabilitation intervention, Figure 1).

The groups were similar on most baseline characteristics, except for a higher proportion of independent walkers (e.g., no use of a walking assistant) in the no physical rehabilitation group, 8.5 % compared to 1.8 % (Table 1).

### Primary outcome: self-reported function (HOOS/KOOS ADL)

#### Analysis: Physical rehabilitation versus no physical rehabilitation

In the primary intention-to-treat analysis for DRAW2, self-reported function measured by the HOOS/KOOS improved from baseline to the 6-week (A1) assessment by 22.8 (95%CI: 21.3 to 24.3) points after physical rehabilitation and by 23.3 (95%CI: 21.2 to 25.5) points after no physical rehabilitation. The difference between groups at the 6-week follow-up was not statistically significant (mean difference of self-reported function: −0.5 points (95% CI: −3.1 to 2.1; p = 0.70). Corresponding changes in self-reported function from baseline to the later follow-ups were: 29.5 (95%CI: 28.0 to 31.1) points for the physical rehabilitation interventions and 28.7 (95%CI: 26.7 to 30.8) points for the no physical rehabilitation comparator at 3 months (A2) and 34.9 (95%CI: 33.3 to 36.5) points for the physical rehabilitation interventions and 34.5 (95%CI: 32.8 to 36.3) points for the no physical rehabilitation comparator at 12 months (A3). The between-group differences were not statistically significant at 3 months (0.8 points: 95%CI: −1.7 to 3.4, p = 0.52) or 12 months (0.4 points: 95%CI: −2.2 to 2.9, p = 0.78).

#### Analysis: home-based telerehabilitation and home-based rehabilitation versus no rehabilitation

The two physical rehabilitation interventions (home-based telerehabilitation and home-based rehabilitation) were compared individually to no physical rehabilitation. The self-reported function improved from baseline to 6-week follow-up (A1) by 24.1 (95%CI: 21.9 to 26.3) points for the home-based telerehabilitation intervention and 21.7 (95%CI: 19.7 to 23.7) points for the home-based rehabilitation intervention. Comparing home-based telerehabilitation to no physical rehabilitation, the between-group difference at 6 weeks (A1) was 0.7 (95%CI: −2.4 to 3.8; p = 0.65) points, while the between-group difference between the home-based rehabilitation intervention and the no physical rehabilitation comparator was −0.1 (95%CI: −3.3 to 3.0; p = 0.94) points at the same time point.

Corresponding changes in self-reported function from baseline to the later follow-ups were: 30.2 (95%CI: 27.9 to 32.4) points for the home-based telerehabilitation intervention and 29.0 (95%CI: 26.9 to 31.1) points for the home-based rehabilitation intervention at 3 months (A2). At 12 months (A3) the improvement was 34.1 (95%CI: 31.5 to 36.7) points for home-based telerehabilitation intervention and 35.6 (95%CI: 3.7 to 37.6) points for the home-based rehabilitation intervention. Comparing home-based telerehabilitation and no physical rehabilitation, the between-group difference was 1.5 (95%CI: −1.6 to 4.5; p = 0.35) points at 3 months (A2) and −0.4 (95%CI: −3.6 to 2.7; p = 0.78) points at 12 months (A3). The between-group difference between home-based rehabilitation intervention and the no physical rehabilitation comparator was 0.3 (95%CI: −2.6 to 3.2; p = 0.85) points at 3 months (A2) and 1.1 (95%CI: −1.6 to 3.7; p = 0.42) points at 12 months (A3).

### Ancillary analyses

The per-protocol analysis, restricted to patients with ≥80% adherence, yielded results consistent with the ITT analysis, confirming no clinically relevant between-group differences at any time point (see supplements, table 3).

#### Harms

During the trial, 13 adverse events occurred in 13 participants (Table 4); three in the home-based telerehabilitation group, six in the home-based rehabilitation group and four in the no physical rehabilitation group.

## DISCUSSION

This replication trial confirmed the main finding from the DRAW1 trial: that physical rehabilitation, after THA and TKA, was not found to be superior to no rehabilitation in terms of self-reported function at 6 weeks (primary end-point, and at 3 and 12 months (secondary endpoints). Importantly, the confidence intervals for both the intention-to-treat and per-protocol analyses excluded the predefined minimal clinically important difference of 10 points (95%CI: −3.1 to 3.4 and −4.0 to 1.6, respectively) at any time point, supporting the robustness of this finding. Thus, the fundamental clinical effectiveness of physical rehabilitation, after THA and TKA, could not be established.

Taken together, the DRAW1 and DRAW2 trials provide remarkably consistent results. In DRAW1, the between-group difference in self-reported function (HOOS/KOOS ADL) at 6 weeks was 3.3 points (95% CI: –1.9 to 8.6; p = 0.10), and in DRAW2, the difference was 0.5 (95%CI: −3.1 to 2.1; p = 0.70). Both trials found confidence intervals that excluded the predefined minimal clinically important difference of 10 points, and both showed a lack of statistically or clinically meaningful superiority of rehabilitation over no rehabilitation. This replication thus strongly reinforces the robustness of the original finding.

Reproducibility is foundational to trustworthy science, particularly for trials with practice-changing implications. As such, the replication of DRAW1 in DRAW2 strengthens the internal validity and credibility of both findings. Replication studies are rare in clinical rehabilitation research, and this trial contributes a rare and methodologically rigorous example—preregistered, adequately powered, and conducted at the same site using the same protocol and population. While small deviations occurred (e.g., split baseline assessments), these are unlikely to have biased the results.

Some may argue that the lack of superiority observed in DRAW1 and DRAW2 is due to suboptimal intervention content—that more intensive or differently structured exercise programs might have produced different results. As researchers with backgrounds in physiotherapy, we acknowledge this perspective and have ourselves previously advocated for more intensive rehabilitation based on earlier assumptions about its efficacy ^31^. However, both trials were deliberately designed as pragmatic and clinically grounded evaluations, using exercise protocols that align closely with current practice and national guidelines for postoperative rehabilitation following THA and TKA ^32–34^. Importantly, no specific rehabilitation protocol has consistently demonstrated superiority over others in systematic reviews and large RCTs ^1,2,35–38^ – including enriched trials in patients at risk for poor post-operative outcome after TKA ^39,40^. This lack of differential effect across intervention types reinforces the notion that the fundamental clinical effectiveness of physical rehabilitation in this population may be limited—at least within the timeframe and outcomes studied. The interventions in DRAW1 and DRAW2 were not chosen because they represented an idealized form of rehabilitation, but because they reflect what is routinely offered in clinical settings, thereby enhancing external validity and decision-making relevance.

In total, 13 adverse events occurred during the trial (three in home-based telerehabilitation group, six in the home-based rehabilitation group, and four in the no physical rehabilitation group) (table 4). Although the number of adverse events differed slightly across groups, there was no apparent clustering, and events were relatively evenly distributed. This suggests that no single rehabilitation approach was associated with a markedly higher risk of adverse events.

Although speculative, the findings from DRAW1 and DRAW2 suggest that natural recovery may account for much of the functional improvement after THA or TKA. This challenges the prevailing assumption that all patients should routinely receive structured rehabilitation. It also resonates strongly with the Choosing Wisely campaign ^9^, which advocates avoiding low-value interventions that offer little benefit relative to their burden on patients and health systems. Applying this principle to postoperative rehabilitation suggests that structured exercise therapy should be offered selectively—ideally through shared decision-making with the patient—and focused on those with the greatest potential to benefit ^41^.

In conclusion, the main finding from the DRAW1 trial was replicated and post-surgical physical rehabilitation following THA or TKA was once not found to be superior to no post-surgical physical rehabilitation in terms of self-reported function in the first year after surgery and rehabilitation. These findings question the default use of postoperative rehabilitation following THA or TKA and instead points towards involving patients in shared-decision making around their needs, options and preferences in relation to post-surgical rehabilitation.

## Supporting information

Figure 1. Flowchart

## Data Availability

All data produced in the present study are available upon reasonable request to the authors

https://dataverse.harvard.edu/dataset.xhtml?persistentId=doi:10.7910/DVN/BUNJQV

## Acknowledgment

The authors would like to thank the Department of Rehabilitation at the Regional Municipality of Bornholm for their invaluable input and support throughout the research process, with special thanks to the involved trial physiotherapists, whose insights, expertise, and dedication made this trial possible. In addition, we would like to extend our sincere gratitude to all participants who generously gave their time and effort to this project. Lastly, we would like to thank the funding parties, the Regional Municipality of Bornholm and the Association of Danish Physiotherapists and Karen Elise Jensens Fond for providing the necessary financial support to which we are deeply grateful.

## Author contribution

All authors meet the International Committee of Medical Journal Editors (ICMJE) criteria for authorship. All authors take responsibility for the integrity of the work as a whole, from inception to finished article. Specific, primary contribution to the following points were as follows: Conception and design (TMC, TB, KT), Analysis and interpretation of the data (TMC, TB, KT, TK), Drafting of the article (TMC, TB, KT, TK), Critical revision of the article for important intellectual content (TMC, TB, KT, TK), Final approval of the article (TMC, TB, KT, TK), Provision of the study patients (TMC), Statistical expertise (TK), Obtaining of funding (TMC, TB), Collection and assembly of data (TMC, TK).

## Data sharing statement

The authors commit to making the relevant anonymized patient-level data available on reasonable request.

