## Supplementary material for "Physical rehabilitation versus no physical rehabilitation after total hip and knee arthroplasty: A replication trial in 169 patients with a 12-month follow-up (DRAW2)": Figure 1. Flowchart

**Assessed for eligibility**
(n=479)

221 THA/258 TKA

**Excluded** (n=93)

- Surgery not due to osteoarthritis (n=11)
- Revision surgery (n=13)
- Postoperative complication (n=33)
- Not able to comply with exercise instructions (n=27)
- Discharged to nursing-home facility and/or receiving in-home rehabilitation by home care (n=9)

**Decline participation** (n=217)

- Wanted to receive some kind of exercise rehabilitation (n=88)
- Did not want to risk receiving home-based telerehabilitation (n=47)
- Did not want exercise rehabilitation (n=6)
- Did not have time to participate (n=10)
- Other reasons (personal reasons, motivation etc.) (n=66)

**Randomized**
(n=169)

Allocated to **home-based telerehabilitation**
(n=52)

Allocated to **home-based rehabilitation**
(n=58)

Allocated to **no physical intervention**
(n=59)

**1. Follow up (6 weeks)**

Analyzed as per ITT-analysis (n=52)[a]
Analyzed as per protocol-analysis (n=47)
Lost to first follow up (n=5)

- Four withdrew consent
- One PJI

Analyzed as per ITT-analysis (n=58)[a]
Analyzed as per protocol-analysis (n=54)
Lost to first follow up (n=4)

- One experienced luxation
- One had general nerve inflammation
- One was prescribed outpatient rehab
- One had a PJI

Analyzed as per ITT-analysis (n=59)[a]
Analyzed as per protocol-analysis (n=48)
Lost to first follow up (n=9)

- Two withdrew consent
- Four were prescribed outpatient rehab
- One experienced a DVT
- One hospitalized due to possible DVT
- One PJI

**2. Follow up (3 months)**

Analyzed as per ITT-analysis (n=52)[a]
Analyzed as per protocol-analysis (n=42)
Lost to second follow up (n=5)

- Two withdrew consent
- Two were prescribed outpatient rehab
- One underwent MUA

Analyzed as per ITT-analysis (n=58)[a]
Analyzed as per protocol-analysis (n=47)
Lost to second follow up (n=7)

- Three withdrew consent
- Three were prescribed outpatient rehab
- One underwent MUA

Analyzed as per ITT-analysis (n=59)[a]
Analyzed as per protocol-analysis (n=42)
Lost to second follow up (n=6)

- Five withdrew consent
- One was prescribed outpatient rehab

**3. Follow up (12 months)**

Analyzed as per ITT-analysis (n=52)[a]
Analyzed as per protocol-analysis (n=40)
Lost to third follow up (n=2)

- One withdrew consent
- One underwent MUA

Analyzed as per ITT-analysis (n=58)[a]
Analyzed as per protocol-analysis (n=45)
Lost to third follow up (n=2)

- One withdrew consent
- One had streptococcus infection that required hospitalization

Analyzed as per ITT-analysis (n=59)[a]
Analyzed as per protocol-analysis (n=38)
Lost to third follow up (n=4)

- Two withdrew consent
- One was prescribed outpatient rehab
- One had a cerebral hemorrhage

[a] Number of imputations in intention- to-treat analysis: 5
