## Supplementary material for "Physical rehabilitation versus no physical rehabilitation after total hip and knee arthroplasty: A replication trial in 169 patients with a 12-month follow-up (DRAW2)": Figures and tables.

### Figure legends and tables

**Picture 1.** Features of home-based telerehabilitation (ICURA)

**Please see attached file**

**Picture 2.** Home-based rehabilitation (Exorlive)

**Please see attached file**

**Figure 1**. CONSORT flow diagram showing participant enrollment, allocation, follow-up, and analysis across the three arms of the DRAW2 trial.

**Please see attached file**

#### Table 1. Baseline characteristics

**Table 1. Baseline characteristics**

| | **Physical rehabilitation[a] (n=110)** | **No physical rehabilitation (n=59)** | **Physical rehabilitation grouped by allocation** | |
|---|---|---|---|---|
| | | | **Home-based telerehabilitation (n=52)** | **Home-based rehabilitation (n=58)** |
| | Mean (SD) | Mean (SD) | Mean (SD) | Mean (SD) |
| THA/TKA | 51 THA /59 TKA | 25 THA /34 TKA | 24 THA /28 TKA | 27 THA /31 TKA |
| Days from surgery to baseline assessment | 8.6 (3.0) | 8.7 (2.9) | 8.6 (2.9) | 8.7 (3.2) |
| Operated knee (left/right) | 53/57 | 29/30 | 25/25 | 28/30 |

PREPRINT

| | | | | |
|---|---|---|---|---|
| Women/men | 65/45 | 39/20 | 28/24 | 37/21 |
| Age, yrs | 68.1 (9.0) | 67.6 (8.1) | 69.6 (7.9) | 66.8 (9.7) |
| Height, cm | 171.0 (8.8) | 170.6 (8.4) | 172.0 (9.3) | 170.1 (8.2) |
| Weight, kg | 88.7 (18.2) | 87.1 (17.5) | 88.1 (17.6) | 89.2 (18.8) |
| BMI (kg/m2) | 30.3 (5.6) | 29.8 (4.9) | 29.7 (5.1) | 30.8 (5.9) |
| **Current use of analgesics, percentage of users [b]** | | | | |
| - Paracetamol, % | 88.3 % | 89.8 % | 91.5 % | 83.8 % |
| - NSAID, % | 41.5 % | 45.1 % | 37.7 % | 41.4 % |
| - Opioids, % | 47.9 % | 45.1 % | 51.5 % | 47.2 % |
| - Neuropathic medicine, % | 3.7 % | 1.7 % | 6.9 % | 2.4 % |
| **Walking aid, percentage of users [b]** | | | | |
| - No walking aids, % | 1.8 % | 8.5 % | 0 % | 3.5 % |
| - Use one elbow stick, % | 22.9 % | 25.4 % | 26.9 % | 19.3 % |
| - Use two elbow sticks, % | 64.2 % | 61.0 % | 59.6 % | 68.4 % |
| - Use a walker, % | 11.0 % | 5.1 % | 13.5 % | 8.8 % |
| **Primary outcome** | | | | |
| HOOS/KOOS ADL subscale, points | 51.8 (17.5) | 55.2 (15.4) | 52.6 (17.9) | 51.1 (17.2) |
| **Secondary outcomes** | | | | |
| HOOS/KOOS-subscales | | | | |
| Symptoms, points | 54.1 (18.1) | 56.5 (18.5) | 55.1 (16.4) | 53.1 (19.5) |

| | | | | |
|---|---|---|---|---|
| Pain, points | 54.7 (19.0) | 55.2 (16.5) | 55.6 (19.0) | 53.9 (19.0) |
| Quality of life, points | 39.4 (15.4) | 43.7 (16.1) | 39.7 (16.0) | 39.2 (14.9) |
| Global assessment, points | 45.8 (16.7) | 46.8 (18.5) | 44.2 (15.9) | 47.2 (17.2) |
| 30-s chair stand test (repetitions) [c] | 0 (IQR: 0 to 0) | 0 (IQR: 0 to 3) | 0 (IQR: 0 to 0) | 0 (IQR: 0 to 0) |
| 4x10m fast-paced walk test (m/s) [c] | 0.7 (IQR: 0.5 to 0.9) | 0.8 (IQR: 0.6 to 1.0) | 0.7 (IQR: 0.5 to 1.0) | 0.7 (IQR: 0.5 to 0.8) |
| Physio-time, minutes | 105.6 (29.1) | 94.4 (26.5) | 109.8 (30.2) | 101.9 (27.6) |

*[a] Physical rehabilitation = home-based telerehabilitation and home-based rehabilitation combined.*
*[b] Frequencies and percentages are based on pooled estimates from the multiple imputation data sets (ITT population).*
*[c] Non-normal distribution and therefore presented as median and interquartile range (IQR).*
*THA: total hip arthroplasty. TKA: total knee arthroplasty. BMI: Body Mass Index. NSAID: non-steroidal anti-inflammatory drug. HOOS: Hip disability and Osteoarthritis Outcome Score. KOOS: Knee injury and Osteoarthritis Outcome Score. ADL: activities of daily living.*

#### Table 2. Changes from baseline to follow ups between physical rehabilitation and no physical rehabilitation

| **Table 2. Changes from baseline to follow ups between physical rehabilitation and no physical rehabilitation** | | | | | |
|---|---|---|---|---|---|
| | Follow up | Physical rehabilitation[a] (95%CI) | No physical rehabilitation (95%CI) | Mean difference (95%CI) | P-value |
| **Primary outcome** | | | | | |
| HOOS/KOOS ADL subscale, points | 6-week follow-up | 22.8 (21.3 to 24.3) | 23.3 (21.2 to 25.5) | -0.5 (-3.1 to 2.1) | 0.70 |
| **Secondary outcome** | | | | | |
| HOOS/KOOS: ADL, points | 3-months follow-up | 29.5 (28.0 to 31.1) | 28.7 (26.7 to 30.8) | 0.8 (-1.7 to 3.4) | 0.52 |

PREPRINT

| | | | | | |
|---|---|---|---|---|---|
| | 12-months follow-up | 34.9 (33.3 to 36.5) | 34.5 (32.8 to 36.3) | 0.4 (-2.2 to 2.9) | 0.78 |
| HOOS/KOOS: Symptoms, points | 6-week follow-up | 12.8 (11.1 to 14.5) | 14.9 (12.7 to 17.0) | -2.1 (-4.9 to 0.7) | 0.15 |
| | 3-months follow-up | 19.8 (18.3 to 21.3) | 18.3 (16.0 to 20.7) | 1.4 (-1.3 to 4.1) | 0.29 |
| | 12 months follow-up | 29.7 (28.1 to 31.4) | 28.2 (26.1 to 30.4) | 1.5 (-1.2 to 4.2) | 0.28 |
| HOOS/KOOS: Pain, points | 6-week follow-up | 17.5 (16.0 to 19.0) | 19.2 (17.6 to 20.8) | -1.7 (-4.1 to 0.6) | 0.15 |
| | 3-months follow-up | 25.1 (23.5 to 26.7) | 26.2 (24.2 to 28.2) | -1.2 (-3.8 to 1.5) | 0.39 |
| | 12-months follow-up | 32.9 (31.2 to 34.5) | 35.0 (33.2 to 36.8) | -2.1 (-4.7 to 0.4) | 0.10 |
| HOOS/KOOS: Quality of life, points | 6-week follow-up | 19.9 (18.2 to 21.5) | 19.0 (17.0 to 20.9) | 0.0 (-1.7 to 3.5) | 0.50 |
| | 3-months follow-up | 28.4 (26.7 to 30.1) | 27.0 (24.9 to 29.1) | 1.44 (-1.3 to 4.2) | 0.30 |
| | 12-months follow-up | 36.9 (35.1 to 38.8) | 36.8 (34.7 to 39.0) | 0.08 (-2.9 to 3.0) | 0.96 |
| Global assessment, points | 6-week follow-up | 22.3 (20.6 to 23.9) | 24.2 (21.9 to 26.4) | -1.9 (-4.7 to 0.9) | 0.17 |
| | 3-months follow-up | 33.2 (31.6 to 34.8) | 33.4 (30.9 to 35.9) | -0.23 (-3.1 to 2.6) | 0.87 |
| | 12-months follow-up | 38.1 (36.3 to 39.8) | 37.3 (34.8 to 39.8) | 0.76 (-2.2 to 3.8) | 0.62 |
| Satisfaction, percentage | 6-week follow-up | 81.0 (79.6 to 82.4) | 80.2 (78.5 to 81.8) | 0.83 (-1.4 to 3.1) | 0.47 |

PREPRINT

| | | | | | |
|---|---|---|---|---|---|
| Exercise adherence, percentage | 6-week follow-up | 80.7 (78.8 to 82.6) | | | |
| 30-s chair stand test (repetitions) [b] | 6-week follow-up | 7 (IQR: 0 to 11) | 6 (IQR: 0 to 11) | | 0.95 |
| | 3-months follow-up | 11 (IQR: 4 to 13) | 9 (IQR: 5 to 12) | | 0.06 |
| | 12-months follow-up | 12 (IQR: 8 to 14) | 10 (IQR: 7 to 16) | | 0.23 |
| 4x10m. fast paced walk test (m/s) [b] | 6-week follow-up | 0.5 (IQR: 0.3 to 0.6) | 0.4 (IQR: 0.3 to 0.6) | | 0.02 |
| | 3-months follow-up | 0.5 (IQR: 0.4 to 0.7) | 0.5 (IQR: 0.3 to 0.7) | | 0.75 |
| | 12-months follow-up | 0.6 (IQR: 0.5 to 0.8) | 0.6 (IQR: 0.4 to 0.9) | | 0.41 |
| Consultation time, (minutes) | 6-week follow-up | 74.1 (72.8 to 75.4) | 71.5 (69.5 to 73.6) | 2.6 (0.2 to 4.9) | 0.03 |
| | 3-months follow-up | 61.4 (60.0 to 62.9) | 65.8 (64.2 to 67.4) | - 4.4 (-6.6 to - 2.1) | 0.00 |
| | 12-months follow-up | 62.7 (61.3 to 64.1) | 58.6 (56.8 to 60.5) | 4.1 (1.8 to 6.4) | 0.00 |
| **Percentage of users of analgesics [c,d]** | | | | | |
| Paracetamol, % | 6-week follow-up | 43.3 % | 52.2 % | | 0.01 |
| | 3-months follow-up | 32.2 % | 25.1 % | | 0.03 |
| | 12-months follow-up | 20.7 % | 18.3 % | | 0.40 |
| NSAID, % | 6-week follow-up | 17.8 % | 14.2 % | | 0.18 |

PREPRINT

| | | | | | |
|---|---|---|---|---|---|
| | 3-months follow-up | 11.5 % | 17.3 % | | 0.02 |
| | 12-months follow-up | 5.8 % | 5.8 % | | 0.97 |
| Opioids, % | 6-week follow-up | 7.3 % | 3.7 % | | 0.04 |
| | 3-months follow-up | 5.8 % | 2.0 % | | 0.01 |
| | 12-months follow-up | 5.5 % | 4.8 % | | 0.66 |
| Neuropathic medicine, % | 6-week follow-up | 2 % | 0 % | | 0.01 |
| | 3-months follow-up | 3.8 % | 0 % | | 0.00 |
| | 12-months follow-up | 5.1 % | 2.0 % | | 0.03 |
| **Percentage of independent walkers [c,d]** | | | | | |
| | 6-week follow-up | 70.2 % | 75.3 % | | 0.00 |
| | 3-months follow-up | 86.7 % | 92.5 % | | 0.00 |
| | 12-months follow-up | 93.1 % | 93.2 % | | 0.10 |

[a] *Physical rehabilitation = home-based telerehabilitation and home-based rehabilitation combined.*

[b] *None-normal distributed, and presented with median and interquartile range (IQR), and compared using the wilcoxon rank sum test.*

[c] *The use of analgesics and numbers of independent walkers were analyzed using the Chi Squared test.*

[d] *Frequencies and percentages are based on pooled estimates from the multiply imputation data sets. NSAID: non-steroidal anti-inflammatory drug. HOOS: Hip disability and Osteoarthritis Outcome Score. KOOS: Knee injury and Osteoarthritis Outcome Score. ADL: activities of daily living.*

*Follow ups. A1: after the 6-week intervention. A2: 3 months postoperatively. A3: 12 months postoperatively.*

PREPRINT

**Table 3. Between-group differences at follow-ups by the individual physical rehabilitations (home-based telerehabilitation and home-based rehabilitation) compared to No physical rehabilitation.**

**Table 3. Comparing the two individual physical rehabilitations interventions to no physical rehabilitation.**

| Outcomes | | Mean difference (95%CI) | P-value | Mean difference (95%CI) | P-value |
|---|---|---|---|---|---|
| **Primary outcome** | | **Home-based telerehabilitation vs. No physical rehabilitation** | | **Home-based rehabilitation vs. No physical rehabilitation** | |
| HOOS/KOOS-ADL, point difference | 6-week follow-up | 0.7 (-2.4 to 3.8) | 0.65 | -0.1 (-3.3 to 3.0) | 0.94 |
| **Secondary outcome** | | | | | |
| HOOS/KOOS-ADL, point difference | 3-months follow-up | 1.5 (-1.6 to 4.5) | 0.35 | 0.3 (-2.6 to 3.2) | 0.85 |
| | 12-months follow-up | -0.4 (-3.6 to 2.7) | 0.78 | 1.1 (-1.6 to 3.7) | 0.42 |
| Symptoms, point difference | 6-week follow-up | -0.5 (-3.7 to 2.8) | 0.77 | -3.5 (-6.7 to -0.3) | 0.03 |
| | 3-months follow-up | 1.4 (-1.9 to 4.7) | 0.40 | 1.5 (-1.6 to 4.6) | 0.35 |
| | 12-months follow-up | 1.9 (-1.2 to 5.0) | 0.22 | 1.1 (-2.1 to 4.3) | 0.49 |
| Pain, point difference | 6-week follow-up | 0.6 (-2.0 to 3.2) | 0.66 | -3.8 (-6.4 to -1.2) | 0.00 |
| | 3-months follow-up | -0.2 (-3.3 to 2.8) | 0.89 | -2.0 (-5.0 to 1.0) | 0.19 |
| | 12-months follow-up | -1.2 (-4.1 to 1.8) | 0.43 | -3.0 (-5.8 to -0.2) | 0.04 |
| Quality of life, point difference | 6-week follow-up | 2.0 (-0.8 to 4.9) | 0.16 | -0.1 (-3.3 to 3.0) | 0.94 |
| | 3-months follow-up | 2.1 (-1.1 to 5.2) | 0.20 | 0.9 (-2.2 to 4.0) | 0.58 |
| | 12-months follow-up | 3.1 (0.1 to 6.4) | 0.06 | -2.7 (-6.0 to 0.7) | 0.12 |
| Global assessment, point difference | 6-week follow-up | 0.4 (-2.7 to 3.5) | 0.82 | -3.9 (-7.2 to 0.7) | 0.02 |
| | 3-months follow-up | 2.5 (0.9 to 5.8) | 0.15 | -2.7 (-6.1 to 0.8) | 0.13 |
| | 12-months follow-up | 1.9 (-1.8 to 5.5) | 0.31 | -0.2 (-3.6 to 3.2) | 0.90 |

PREPRINT

| | | | | | |
|---|---|---|---|---|---|
| Satisfaction, point difference | 6-week follow-up | 4.3 (2.0 to 6.6) | 0.00 | -2.3 (5.0 to 0.4) | 0.10 |
| 30-s chair stand test (repetitions) [a] | 6-week follow-up | 8 (IQR: 0 to 11) vs. 6 (IQR: 0 to 11) | 0.10 | 5 (IQR: 0 to 11) vs. 6 (IQR: 0 to 11) | 0.15 |
| | 3-months follow-up | 11 (IQR: 5 to 13) vs. 9 (IQR: 5 to 12) | 0.01 | 10 (IQR: 1 to 13) vs. 9 (IQR: 5 to 12) | 0.42 |
| | 12-months follow-up | 12 (IQR: 7 to 14) vs. 10 (IQR: 7 to 15) | 0.40 | 12 (IQR: 8 to 14) vs. 10 (IQR: 7 to 15) | 0.22 |
| 4x10m fast-paced walk test (m/s) [a] | 6-week follow-up | 0.4 (IQR: 0.3 to 0.6) vs. 0.4 (IQR: 0.3 to 0.6) | 0.06 | 0.4 (IQR: 0.3 to 0.6) vs. 0.4 (IQR: 0.3 to 0.6) | 0.02 |
| | 3-months follow-up | 0.5 (IQR: 0.4 to 0.7) vs. 0.5 (IQR: 0.3 to 0.7) | 0.89 | 0.5 (IQR: 0.3 to 0.7) vs. 0.5 (IQR: 0.3 to 0.7) | 0.68 |
| | 12-months follow-up | 0.6 (IQR: 0.5 to 0.8) vs. 0.6 (IQR: 0.4 to 0.9) | 0.93 | 0.6 (IQR: 0.4 to 0.9) vs. 0.6 (IQR: 0.4 to 0.9) | 0.20 |
| Consultation time difference (minutes) | 6-week follow-up | 1.4 (1.2 to 1.6) | 0.23 | 3.3 (0.6 to 6.0) | 0.02 |
| | 3-months follow-up | -6.6 (-9.1 to -4.1) | 0.00 | -2.4 (-4.9 to 0.2) | 0.07 |
| | 12-months follow-up | 4.2 (2.0 to 7.4) | 0.00 | 3.5 (0.9 to 6.2) | 0.01 |
| **Percentage of users of analgesics [b,c]** | | | | | |
| Paracetamol, % | 6-week follow-up | 39.6 % vs. 52.2 % | 0.00 | 46.6 % vs. 52.2 % | 0.17 |
| | 3-months follow-up | 26.2 % vs. 25.1 % | 0.77 | 37.6 % vs. 25.1 % | 0.00 |
| | 12-months follow-up | 20.4 % vs. 18.3 % | 0.54 | 21.0 % vs. 18.3 % | 0.41 |
| NSAID, % | 6-week follow-up | 8.1 % vs. 14.2 % | 0.02 | 26.6 % vs. 14.2 % | 0.00 |
| | 3-months follow-up | 5.8 % vs. 1.3 % | 0.00 | 16.6 % vs. 17.3 % | 0.81 |
| | 12-months follow-up | 5.8 % vs. 5.8 % | 1.00 | 5.9 % vs. 5.8 % | 1.00 |

PREPRINT

| | | | | | |
|---|---|---|---|---|---|
| Opioids, % | 6-week follow-up | 8.9 % vs. 3.7 % | 0.01 | 5.9 % vs. 3.7 % | 0.23 |
| | 3-months follow-up | 4.6 % vs. 2.0 % | 0.09 | 6.9 % vs. 2.0 % | 0.00 |
| | 12-months follow-up | 4.2 % vs. 4.8 % | 0.77 | 6.6 % to 4.8 % | 0.34 |
| Neuropathic medicine, % | 6-week follow-up | 4.2 % vs. 0.0 % | 0.00 | 0.0 % vs. 0.0 % | ! |
| | 3-months follow-up | 8.1 % vs. 0.0 % | 0.00 | 0.0 % vs. 0.0 % | ! |
| | 12-months follow-up | 6.2 % vs. 2.0 % | 0.01 | 4.1 % vs. 2.0 % | 0.14 |
| **Percentage of independent walkers** | | | | | |
| | 6-week follow-up | 72.7 % vs. 75,3 % | 0.00 | 67.9 % vs. 75.3 % | 0.01 |
| | 3-months follow-up | 86.5 % vs. 92.5 % | 0.00 | 86.9 % vs. 92.5 % | 0.00 |
| | 12-months follow-up | 95.8 % vs. 93.2 % | 0.38 | 90.7 % vs. 93.2 % | 0.00 |

[a]30s chair stand test and 4x10m fast-paced walk test were analysed with Wilcoxon sum rank-test.

[b]*The use of analgesics and use of walking aid were analysed using the Chi Squared test.*

[c]*Frequencies and percentages are based on pooled estimates from the multiply imputation data sets.*

*! = can not be calculated due to no events.*

#### Table 4. Adverse events

Table 4. Adverse events

| Adverse events | Home-based telerehabilitation | Home-based rehabilitation | No physical rehabilitation |
|---|---|---|---|
| Before first follow up | N = 1<br>One had PJI | N = 4<br>One had PJI | N = 3<br>One had PJI |

PREPRINT

| | | | |
|---|---|---|---|
| (6 weeks) | | One had a dislocated hip<br>One had a general nerve inflammation<br>One was hospitalized due to suspected DVT | One was a deep vein thrombosis<br>One was hospitalized due to suspected DVT |
| Before second follow up<br>(3 months) | N = 1<br>One had MUA | N = 1<br>One had MUA | N = 0 |
| Before third follow up<br>(12 months) | N = 1<br>One had MUA | N = 1<br>One had a general streptococcus infection | N = 1<br>One had a cerebral hemorrhage |
| **Total** | **N = 3** | **N = 6** | **N = 4** |

**PJI: periprosthetic joint infection. MUA: manipulation under anesthesia**

PREPRINT
