## Supplements for "Physical rehabilitation versus no physical rehabilitation after total hip and knee arthroplasty: A replication trial in 169 patients with a 12-month follow-up (DRAW2)"

### Supplements for DRAW2

Supplement Table 1. **Outcome assessments at each time point.**

Supplement Table 2. **Comparing exercise adherence between the physical rehabilitation groups**

Supplement Table 3**. Per protocol analysis for exercise adherence**

Supplement Methods. **Multiple imputation analysis**

PREPRINT

| **Supplement Table 1. Outcome data for the three interventions** | | | | |
|---|---|---|---|---|
| | Follow up | Home-based **tele**rehabilitation (SD) | Home-based rehabilitation (SD) | No physical rehabilitation (SD) |
| Self-reported | | | | |
| HOOS/KOOS: ADL, points | Baseline | 52.6 (17.9) | 51.1 (17.2) | 55.2 (15.4) |
| | 6-weeks follow-up | 76.6 (15.8) | 72.8 (17.6) | 78.5 (14.6) |
| | 3-months follow-up | 82.7 (13.8) | 80.1 (16.0) | 83.8 (13.1) |
| | 12-months follow-up | 86.5 (13.0) | 86.6 (13.0) | 89.4 (10.8) |
| HOOS/KOOS: symptoms, points | Baseline | 55.1 (16.4) | 53.1 (19.5) | 56.5 (18.5) |
| | 6-weeks follow-up | 69.5 (17.5) | 64.5 (19.1) | 71.4 (17.8) |
| | 3-months follow-up | 74.9 (14.4) | 72.9 (16.2) | 74.9 (17.4) |
| | 12-months follow-up | 85.3 (12.0) | 82.5 (14.4) | 84.8 (14.1) |
| HOOS/KOOS: pain, points | Baseline | 55.6 (19.0) | 53.9 (19.0) | 55.2 (16.5) |
| | 6-weeks follow-up | 75.4 (17.6) | 69.3 (18.8) | 74.5 (17.1) |
| | 3-months follow-up | 81.6 (15.9) | 78.1 (18.2) | 81.5 (16.8) |
| | 12-months follow-up | 89.4 (11.7) | 85.9 (13.7) | 90.2 (12.3) |
| HOOS/KOOS: quality of life, points | Baseline | 39.7 (16.0) | 39.2 (14.9) | 43.7 (16.1) |
| | 6-weeks follow-up | 60.7 (19.2) | 58.1 (21.6) | 62.7 (19.7) |
| | 3-months follow-up | 68.7 (20.2) | 67.1 (21.1) | 70.7 (19.8) |
| | 12-months follow-up | 79.7 (19.8) | 73.4 (22.4) | 80.6 (18.5) |
| Global assessment | Baseline | 44.2 (15.9) | 47.2 (17.2) | 46.8 (18.5) |
| | 6-weeks follow-up | 68.7 (15.2) | 67.5 (20.4) | 71.0 (16.1) |
| | 3-months follow-up | 80.1 (14.0) | 78.0 (16.1) | 80.2 (15.5) |
| | 12-months follow-up | 83.4 (16.8) | 84.3 (15.2) | 84.1 (17.2) |
| Satisfaction, % | 6-weeks follow-up | 84.5 (13.6) | 77.9 (18.7) | 80.2 (14.4) |
| Exercise adherence, % | 6-weeks follow-up | 82.5 (21.2) | 79.0 (23.7) | N/A |
| Performance-based | | | | |
| 30s chair stand test (repetitions) | Baseline | 0 (IQR: 0 to 0) | 0 (IQR: 0 to 0) | 0 (IQR: 0 to 3) |
| | 6-weeks follow-up | 10 (IQR: 0 to 12) | 9 (IQR: 0 to 12) | 10 (IQR: 0 to 13) |
| | 3-months follow-up | 11 (IQR: 7 to 14) | 12 (IQR: 6 to 13) | 11 (IQR: 9 to 14) |
| | 12-months follow-up | 12 (IQR: 9 to 15) | 13 (IQR: 10 to 15) | 13 (IQR: 10 to 16) |
| 4x10m. fast paced walk test (m/s) | Baseline | 0.7 (IQR: 0.5 to 1.0) | 0.7 (IQR: 0.5 to 0.8) | 0.8 (IQR: 0.6 to 1.0) |
| | 6-weeks follow-up | 1.2 (IQR: 1.0 to 1.4) | 1.2 (IQR: 0.9 to 1.3) | 1.3 (IQR: 1.0 to 1.5) |
| | 3-months follow-up | 1.3 (IQR: 1.1 to 1.5) | 1.3 (IQR: 1.1 to 1.4) | 1.4 (IQR: 1.2 to 1.5) |
| | 12-months follow-up | 1.4 (IQR: 1.2 to 1.6) | 1.4 (IQR: 1.2 to 1.6) | 1.5 (IQR: 1.2 to 1.6) |
| Consultation time (minutes) | Baseline | 109.8 (30.2) | 101.9 (27.6) | 94.4 (26.5) |
| | 6-weeks follow-up | 73.3 (15.7) | 74.8 (15.5) | 71.5 (17.9) |
| | 3-months follow-up | 59.2 (15.9) | 63.5 (17.4) | 65.8 (14.2) |
| | 12-months follow-up | 63.3 (16.1) | 62.2 (16.2) | 58.6 (16.3) |

| **Supplement Table 2. Comparison of exercise adherence between the physical rehabilitation groups** | | | | | |
|---|---|---|---|---|---|
| | Follow up | Home-based telerehabilitation | Homebased rehabilitation | Mean difference (95%CI) | P-value |
| Exercise adherence, percentage | 6-weeks follow-up | 82.5 (79.9 to 85.1) | 79.0 (76.2 to 81.7) | 3.6 (-0.2 to 7.3) | 0.07 |

*The two-sample t test were used comparing exercise adherence between the two physical rehabilitation groups.*

| Supplement Table 3. Per protocol analysis for exercise adherence | | | | |
|---|---|---|---|---|
| | **Physical rehabilitation** | **No physical rehabilitation** | Between-group difference | |
| HOOS/KOOS: ADL, difference from baseline | Mean difference (95%CI) | Mean difference (95%CI) | Mean difference (95%CI) | P-value |
| 6-weeks follow-up | 22.0 (20.4 to 23.7) | 23.2 (20.9 to 25.5) | -1.2 (-4.0 to 1.6) | 0.40 |
| 3-months follow-up | 29.1 (27.3 to 30.9) | 28.3 (26.1 to 30.5) | 0.8 (-2.0 to 3.6) | 0.57 |
| 12-months follow-up | 33.8 (31.9 to 35.8) | 34.4 (32.5 to 36.2) | -0.5 (-3.3 to 2.3) | 0.71 |
| 75 out of 110 (68 %) patients allocated to physical rehabilitation reported their exercise adherence to 80 % or above, and were included in the per protocol analysis. | | | | |

#### Supplement Methods. Multiple imputation analysis.

Five imputed datasets were created in the analysis. The extent of missing data per variable were as follows:

**At baseline, A0**: height: 2 missing, weight: 3 missing, paracetamol: 7 missing, NSIAD: 7 missing, Opioid: 7 missing, neuropathic: 7 missing, self-reported symptoms: 5 missing, self-reported pain: 5 missing, self-reported function: 5 missing, self-reported QoL: 5 missing, self-reported global assessment: 5 missing, 4x10m. fast walking test: 1 missing, time: 7 missing. **At first follow-up, A1**: self-reported symptoms: 22 missing, self-reported pain: 22 missing, self-reported function: 22 missing, self-reported quality of life: 21 missing, self-reported global assessment: 22 missing, satisfaction: 25 missing, exercise adherence: 25 missing, 30s chair-stand test: 11 missing, 4x10m. fast walking test: 12 missing, paracetamol: 11 missing, NSAID: 11 missing, opioids: 11 missing, neuropathic: 11 missing, walking aid: 11 missing, time: 28 missing. **At second follow-up, A2**: self-reported symptoms: 38 missing, self-reported pain: 37 missing, self-reported function: 37 missing, self-reported quality of life: 36 missing, self-reported global assessment: 36 missing, 30s chair-stand test: 22 missing, 4x10m. fast walking test: 22 missing, paracetamol: 19 missing, NSAID: 19 missing, opioids: 19 missing, neuropathic: 19 missing, walking aid: 24 missing, time: 40 missing. **At third and final follow-up, A3**: self-reported symptoms: 45 missing, self-reported pain: 45 missing, self-reported function: 45 missing, self-reported quality of life: 45 missing, self-reported global assessment: 45 missing, 30s chair-stand test: 47 missing, 4x10m. fast walking test: 46 missing, paracetamol: 46 missing, NSAID: 46 missing, opioids: 46 missing, neuropathic: 46 missing, walking aid: 45 missing, time: 46 missing.

The specific imputation models were fitted for each missing variable, models included type of surgery, gender, age, and prior assessment of the specific outcomes.

PREPRINT
